# Accurate Classification of the Gut Microbiota of Patients in Intensive Care Units During the Development of Sepsis and Septic Shock

**DOI:** 10.1101/2020.02.27.20028761

**Authors:** Wanglin Liu, Mingyue Cheng, Jinman Li, Peng Zhang, Hang Fan, Qinghe Hu, Maozhen Han, Longxiang Su, Huaiwu He, Yigang Tong, Kang Ning, Yun Long

**Author notes:** Equal contribution (Wanglin Liu, Mingyue Cheng, Jinman Li). Corresponding authors. (Tong Y), (Ning K), (Long Y).

## Abstract

The gut microbiota of intensive care unit (ICU) patients display extreme dysbiosis, which is associated with increased susceptibility to organ failure, sepsis and septic shock. However, this dysbiosis is hard to be characterized for each patient, owing to the highly dimensional complexity of gut microbiota. We thus tested whether the concept of enterotype can be applied to the gut microbiota of ICU patients, to describe the dysbiosis. We collected 131 fecal samples from a cohort of 64 ICU patients diagnosed with sepsis or septic shock, using 16S rRNA gene sequencing to dissect their gut microbiota compositions. We find that during the development of sepsis or septic shock, as well as various medical treatments, ICU patients always contain two patterns of dysbiotic microbiota named as ICU-enterotypes, which cannot be explained by the individual host properties such as age, gender and body mass index, as well as external stressors such as infection sites and antibiotic use. ICU-enterotype I comprised predominantly *Bacteroides* and an unclassified genus of family *Enterobacteriaceae*, while ICU-enterotype II comprised predominantly *Enterococcus*. Among more critically ill patients with acute physiology and chronic health evaluation (APACHE) II score > 18, samples of septic shock were more likely to present with ICU-enterotype I (*p* = 0.041). Additionally, ICU-enterotype I was correlated with high serum lactate level (*p* = 0.007). Therefore, different patterns of dysbiosis are correlated with different clinical outcomes, suggesting that the diagnosis of ICU-enterotypes as an independent clinical index is crucial. For this purpose, the microbial-based human index (MHI) classifier we proposed shows high precision and effectiveness in timely monitoring of ICU-enterotypes of an individual patient. Together, our work serves as the first step toward precision medicine for septic patients based on the gut microbiota profile.

## Introduction

In intensive care units (ICUs), patients are in critically ill conditions, along with inflammation and suppression of the immune system. These patients usually undergo frequent medical interventions including administration of antibiotics, vasoactive agents, and opioids. Together, these factors can lead to disruption of the gut microbiota of ICU patients and such dysbiosis may increase susceptibility to hospital-acquired infections, sepsis and multi-organ dysfunction syndrome in return [1–3]. Therefore, the gut microbiota should be carefully treated when performing medical interventions.

To date, a number of preliminary studies have investigated the gut microbiota of ICU patients (shortened as ICU gut microbiota) [4–7]. ICU gut microbiota has been previously characterized as displaying extreme dysbiosis, with loss of health-promoting commensal microbes such as those belonging to the phyla Firmicutes and Bacteroidetes, and an increase of pathogenic microbes such as Proteobacteria [6]. However, this dysbiosis is hard to be characterized for each patient, owing to the high heterogeneity of gut microbiota. Thus, it is necessary for researchers to find a proper way to characterize the ICU gut microbiota, which is meaningful to clinical diagnosis.

Recently, enterotype analysis [8,9] has been applied to a variety of studies referring to gut microbiota, especially the enterotypes of healthy human gut (termed as conventional enterotypes). It is interesting that a certain number of studies have observed two patterns of conventional enterotypes [10,11], in spite of the distinction of diet, genetic materials and environmental exposure among those studies. Therefore, we hypothesize that two or more patterns of ICU gut microbiota (termed as ICU-enterotypes) can be observed in ICU patients, in spite of the distinction of the inherited traits and external stressors such as the infection types and antibiotic use.

In this study, we recruited a cohort of ICU patients with sepsis or septic shock to confirm our hypothesis, through investigating their gut microbiota composition. We observed that two patterns of ICU-enterotypes were pervasive during the nine-day sampling, even though the subjects were of quite heterogeneity in the infection types and antibiotic use. Moreover, we found patients with septic shock were more likely to present with ICU-enterotype I, which also correlated with high serum lactate level. These results might be caused by the microbiota patterns of ICU-enterotype I, which contained dominant unclassified genus of family *Enterobacteriaceae* and *Bacteroides*, compared to ICU-enterotype II which contained predominantly *Enterococcus*. Additionally, the MHI classifier proposed in this study can facilitate timely monitoring of the ICU-enterotypes of patients towards microbiome-based precision medicine.

## Results

A cohort of 64 ICU patients (Aged 57.71 ± 18.85) who developed sepsis or septic shock were enrolled in this study. Thirty-two patients received carbapenem administration with or without combination of other types of antibiotics (classified as carbapenem use group), while 30 patients received other types of antibiotics such as cephalosporin, penicillin, azithromycin, fluoroquinolones or vancomycin (classified as non-carbapenem use group). Patients received carbapenem were separated because of its broad-spectrum activity against bacteria and its wide use in critically ill patients [12,13]. The remaining two patients did not use antibiotics during sample collecting duration. We collected 131 fecal samples from these 64 patients during their first nine days in ICU for ICU-enterotype identification. To avoid repeated measures from the same individual, we then used their first fecal samples (n = 64) and the corresponding clinical information, such as sequential organ failure assessment (SOFA) score (10.67 ± 4.02), acute physiology and chronic health evaluation (APACHE) II score (20.37 ± 8.14), and lactate level (2.33 ± 2.88), to investigate on the correlation between microbiome compositions and clinical parameters. The antibiotic usage and the detailed clinical information of 64 patients at the day of the collection of their first fecal samples are available in Table S1. The details of the distribution and characteristics of the 131 fecal samples obtained over the nine days are available in Figure S1 and Table S2.

### Two ICU-enterotypes were identified in ICU gut microbiota

Two ICU-enterotypes were identified using the same methods reported in the original paper, with qualified statistical evaluation (Table S3). The distributions of taxa at phylum, class, order, family, genus levels showed significant differences in two ICU-enterotypes (Table S4). The microbial richness (quantified by the number of observed operational taxonomic units (OTUs)) was lower in samples of ICU-enterotype I than samples in ICU-enterotype II (*p* = 2.064e-4, Mann-Whitney-Wilcoxon Test), while the microbial diversity (Shannon index) showed no statistically significant difference (*p* = 0.05729). As shown in **Figure 1**A and B, at phylum level, Bacteroidetes were more prevalent in ICU-enterotype I (*p* = 3.08607e-14), while Actinobacteria and Firmicutes were more prevalent in ICU-enterotype II (*p* = 6.19996e-10, *p* = 9.24951e-08, Kruskal-Wallis test, respectively). At genus level, *Bacteroides* and *Parabacteroides* were more prevalent in ICU-enterotype I (*p* = 6.65034e-12, *p* =1.48078e-11, respectively), while *Enterococcus, Vagococcus* and *Lactobacillus* were more prevalent in ICU-enterotype II (*p* = 7.51562e-13, *p* = 2.14396e-11, *p* = 0.001992467, respectively). Importantly, an unclassified genus of the family *Enterobacteriaceae* was quite dominant in ICU-enterotype I (0.251 ± 0.257 in ICU-enterotype I, 0.082 ± 0.0165 in ICU-enterotype II, *p* = 9.49e-06).

**Figure 1.**
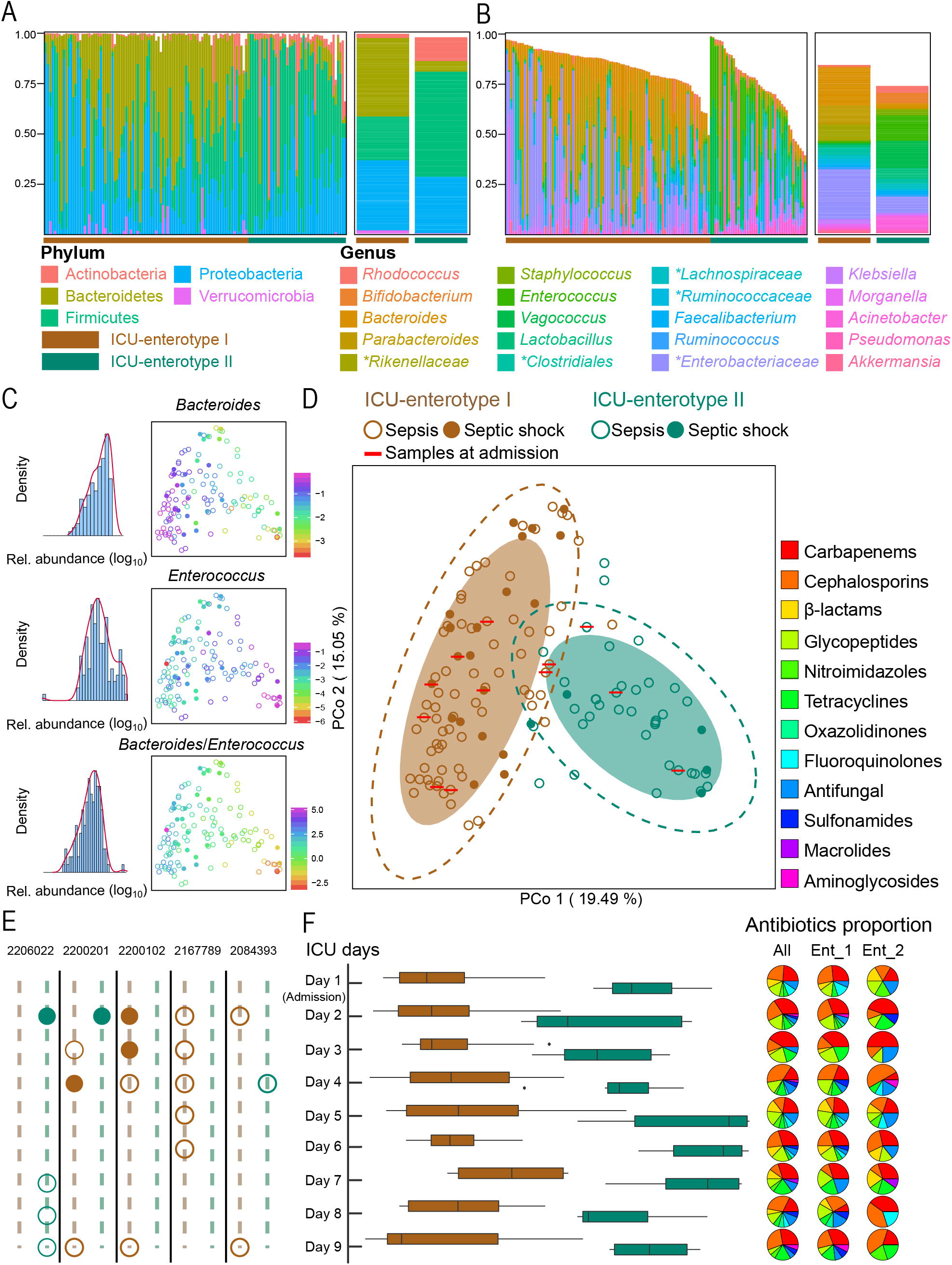
Two ICU-enterotypes are identified in ICU patients with sepsis and septic shock. **A** and **B**. Taxonomic composition of 131 fecal samples at phylum (**A**) and genus (**B**) level. Taxa whose relative abundance > 1% among all samples are plotted. Unclassified genera are designated as a higher rank marked by asterisks. The left panel represents the taxonomic composition of each sample, while the right panel represents the taxonomic composition of two ICU-enterotypes using mean abundance calculated from the data in left panel. The brown and green bars represent the ICU-enterotypes to which these samples belong. **C**. Distributions of the log-transformed (log10) relative abundance of the most significantly differing genera. The left panel displays the observed distributions using frequency distribution histogram with density curve. The right panel displays these distributions in ICU-enterotype space represented by Jensen–Shannon distance (JSD)-based principal coordinate analysis plot (PCoA) plot. The solid circles refer to samples of septic shock, while the hollow circles refer to samples of sepsis. Color in PCoA plot: log-transformed (log10) relative abundances of the genera for each sample. **D**. Gut microbiota compositions of individual patients in ICU-enterotype I (n = 89) and ICU-enterotype II (n= 42) are plotted on a JSD-based PCoA plot at genus level. Shaded ellipses represent the 80% confidence interval, while the dotted ellipse borders represent the 95% confidence interval. **E**. The variation of ICU-enterotypes of five individual patients (with their ID shown at the top) along the nine-day observation. Results for other patients are shown in Figure S1. **F**. In the left panel, the distributions of fecal samples on each day are plotted against the PCo1 axis of (D). Boxes represent the interquartile range (IQR) between first and third quartiles and the line inside represents the median. Whiskers denote the lowest and highest values within 1.5 × IQR from the first and third quartiles, respectively. In the right panel, the proportions of antibiotics used on each day are displayed using all samples, samples of ICU-enterotype I, ICU-enterotype II, respectively.

As shown in Figure 1C, two most discriminating genera named *Bacteroides* (dominant in ICU-enterotype I) and *Enterococcus* (dominant in ICU-enterotype II), as well as their ratio, showed obvious gradient distributions against the PCo 1 axis. Both these distributions were close to log-normal, suggesting the enrichment of *Bacteroides* in ICU-enterotype I and *Enterococcus* in ICU-enterotype II. These results indicate that, unlike the conventional enterotypes characterized by distributions of *Bacteroides and Prevotella* [8], ICU-enterotypes largely depend on the distributions of *Bacteroides* and *Enterococcus*. Moreover, the gradient distributions of the unclassified genus of the family *Enterobacteriaceae* was observed against PCo 2 axis (Figure S2). Considering its dominant abundance in ICU-enterotype I and potential pathogenicity, its identity deserves further investigations.

We found that the pathogens identified from positive bacterial culture tests of the infection sites of patients partially overlapped with the gut microbiome profiles at genus level (Table S1, Table S4). Most of the overlapped genera such as *Enterococcus, Staphylococcus, Corynebacterium* and *Acinetobacter* were observed differently distributed between two ICU-enterotypes. It confirmed the associations between gut microbiome and causative pathogens outside the intestine.

### ICU-enterotypes are pervasive in the cohort of ICU patients

The two identified ICU-enterotypes are pervasive in the cohort of ICU patients with sepsis and septic shock. As was shown in Figure 1D, the samples of admission mixed within all samples collected during ICU days, without any clustering pattern. Additionally, they were also distributed among two ICU-enterotypes. We then displayed the distributions of all 131 samples on each of nine days to test whether the enterotypes remained when using one single sample of different patients (Figure 1F). Interestingly, the ICU-enterotypes were observed obvious on each day, in spite of the differences in the subject sets and the proportion of various antibiotic categories on each day. We also found that the distribution of carbapenem use before or after ICU admission (Figure S3A and B) showed no significant difference in ICU-enterotypes (Mann-Whitney-Wilcoxon test). The distribution of infection sites was observed to correlate with ICU-enterotype: Samples of non-pulmonary infection enriched in the ICU-enterotype I (Figure S3D). This correlation still needs further validation on a larger scale. We did not deny that the antibiotics and infection types can impact on gut microbiota of ICU patients, however, they were not the determinants of ICU-enterotypes. ICU-enterotypes might be two resulted patterns of the dysbiosis of ICU gut microbiota, caused by intricate factors such as inherited traits, various medical treatments and infection types of ICU patients.

The stability of ICU-enterotypes of individual patients during the first nine days remained unknown, owing to the lack of samples at several time points. ICU-enterotype variation was observed in 14 patients (Figure S1), while no change of ICU-enterotypes were observed for the remaining patients, which was probably due to the small sample size. Notably, ICU-enterotype of a patient (with ID 2167789) was observed quite stable, even on a continuous sampling of five days (Figure 1E). The stability of ICU-enterotypes might be less than the conventional enterotypes, on account of that the ICU patients were frequently exposed to various external stressors that would largely affect the gut microbiota. Considering the relative instability of ICU-enterotypes and their correlation with septic shock observed in the following sections, it is important to realize the timely monitoring of ICU-enterotypes of patients.

### Highly effective binary classifier for discriminating ICU-enterotypes

Identifying the ICU-enterotype of a single sample can facilitate the timely monitoring of ICU-enterotypes. Thus, we proposed a binary classifier for this intention, based on the non-redundant taxonomic biomarkers of ICU-enterotypes, selected by LEfSe and mRMR. As shown in **Figure 2**A and B, ten selected taxonomic biomarkers at different phylogenetic levels were of great discriminant ability with LDA scores (log10) > 4.0, as well as of large differences in abundance proportion in two ICU-enterotypes. Additionally, Bacteroidetes, Firmicutes at phylum level, and *Bacteroides, Enterococcus* at genus level were the most discriminant biomarkers.

**Figure 2.**
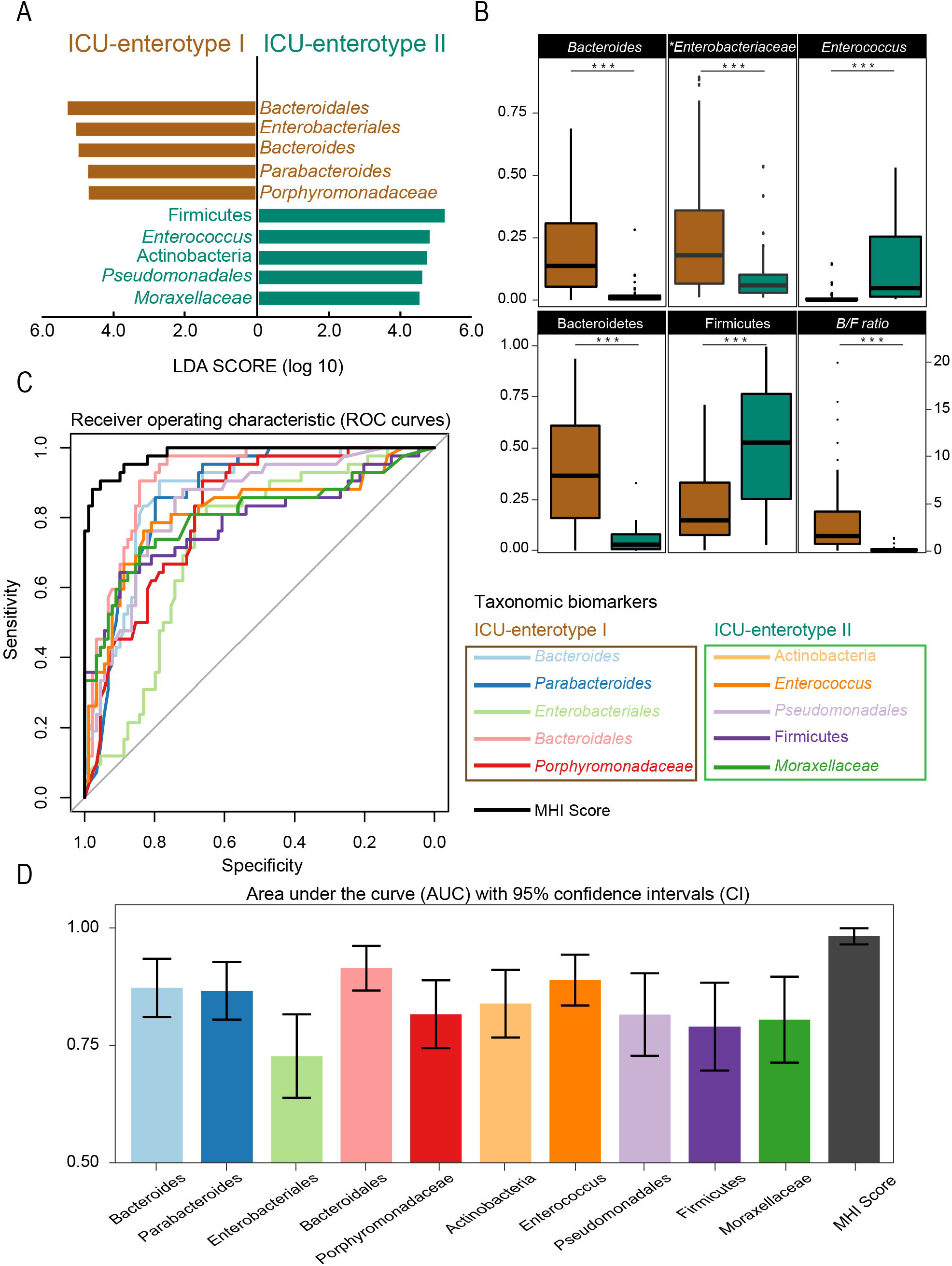
MHI score shows greater discriminant ability than single taxonomic biomarker in ICU-enterotypes classification. **A**. Ten taxonomic biomarkers selected by LEfSe and mRMR are shown with the length of the bar representing the LDA score (log10) and the color indicating which ICU-enterotypes these biomarkers belong to. **B**. The relative abundances of the most dominant biomarkers at phylum and genus level differ largely between two ICU-enterotypes. Boxes represent the interquartile range (IQR) between first and third quartiles and the line inside represents the median. Whiskers denote the lowest and highest values within 1.5 × IQR from the first and third quartiles, respectively. Statistical significance is tested using the Mann–Whitney–Wilcoxon test, ***P < 0.001, n.s., not significant. **C**. T**he receiver operating characteristic (ROC)** is performed for all 131 samples using 10 individual taxonomic biomarkers or the m**icrobial-based human index (MHI)** score. ROC with 95% c**onfident interval (CI) o**f each biomarker or MHI score is displayed in Figure S4. **D**. AUC with 95% CI calculated from the results of (C).

We then combined these ten biomarkers to produce MHI score of each fecal sample, as described in the methods section. We firstly compared the ability of the relative abundance of each individual biomarker and the MHI score, to classify all 131 samples into two ICU-enterotypes (Figure 2C and D, Figure S4). The results showed that the microbial-based human index (MHI) score improved effectiveness of classifying these samples into ICU-enterotype I or II, with the highest area under the curve (AUC) value of 0.9791 (95% CI: 0.9586–0.9997) compared with any individual taxonomic biomarker.

We then proposed a threshold of MHI score as the judgment criteria of the MHI classifier for convenient clinical diagnosis. The threshold was trained to 1.017 in the training set of 106 samples and then applied to the testing set of 25 samples. As shown in **Table 1**, the classifier with this trained threshold (1.017) showed an AUC of 0.982 and an F1 score of 0.929 during the training process. Moreover, this classifier was effective for classification during the testing process as well, with an AUC of 0.985 and an F1 score of 0.903. Taken together, these results indicate that the combination of these ten taxonomic biomarkers as the MHI score has the potential to serve as an effective classification strategy for assessing ICU-enterotypes in ICU clinical trial. Its practical performance on other septic cohort in ICU clinical trials still needs further evaluation.

**Table 1.**
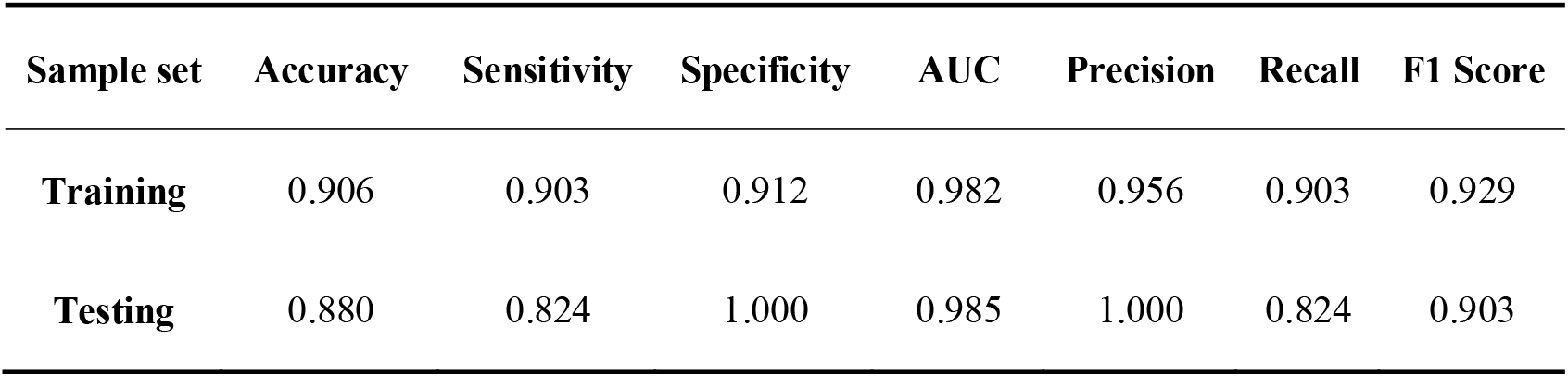
Evaluation of the classification ability of MHI classifier.

| Sample set | Accuracy | Sensitivity | Specificity | AUC | Precision | Recall | F1 Score |
| --- | --- | --- | --- | --- | --- | --- | --- |
| Training | 0.906 | 0.903 | 0.912 | 0.982 | 0.956 | 0.903 | 0.929 |
| Testing | 0.880 | 0.824 | 1.000 | 0.985 | 1.000 | 0.824 | 0.903 |

### Two ICU-enterotypes are correlated with septic shock and serum lactate

#### Correlation with septic shock

The ICU-enterotypes were observed to correlate with septic shock. As shown in Figure 1D and **Table 2**, the samples of sepsis (106 samples from 46 patients) were highly prevalent in both ICU-enterotype I (69 samples from 33 patients, 65.1%) and ICU-enterotype II (37 samples from 13 patients, 34.9%). However, the samples of septic shock (25 samples from 18 patients) were mostly classified as ICU-enterotype I (20 samples from 14 patients, 80%), with a small number classified as ICU-enterotype II (five samples from four patients, 20%). Considering the microbiome compositions and clinical conditions (such as the status of sepsis or septic shock and APACHE II score) of each sample varied every day under clinical intervention, we then viewed these samples as independent. Higher APACHE II score indicates severer disease condition [14–16] and we found that in this study, patients who did not survive had higher APACHE II score than those who survived (median ± interquartile range: 18.5 ± 11 versus 22.5 ± 14, *p* = 0.1, Mann–Whitney–Wilcoxon test), though non-statistically significant. We thus set a cutoff value for APACHE II score as > 18 to decide whether samples are from patients with severer disease conditions. Among samples with an APACHE II score > 18 (83 out of 131 samples), a significantly larger proportion (86.7% vs. 57.4%, *p* = 0.041, Fisher’s exact test) of samples of septic shock presented with ICU-enterotype I than that of samples of sepsis. Nevertheless, among samples with an APACHE II score ≤ 18, no significant difference (*p* = 0.675) between sepsis and septic shock was observed. These results indicate that patients with septic shock are more likely to present with ICU-enterotype I, especially in the case of a more critical status.

**Table 2.**
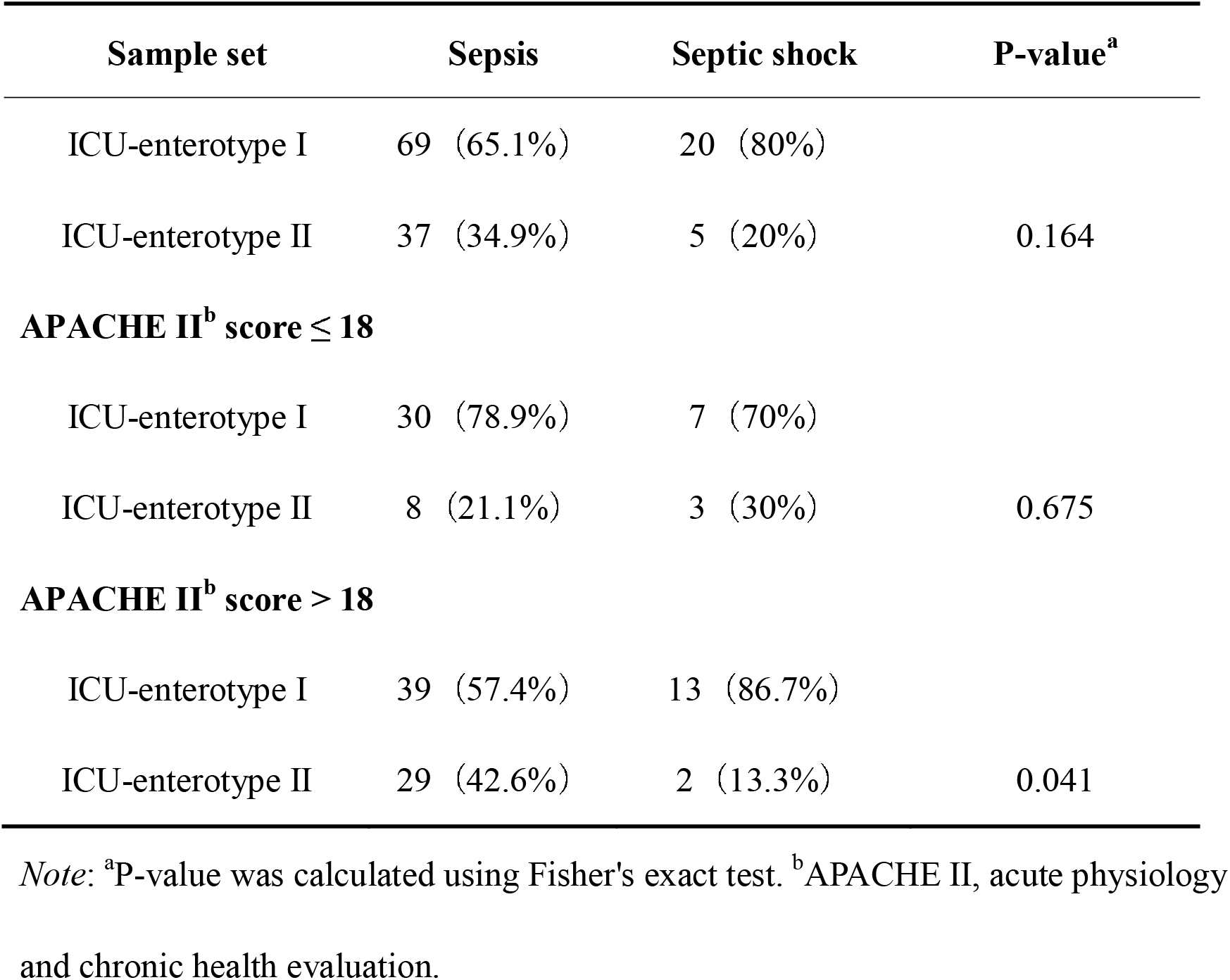
Distributions of ICU-enterotypes differ between patients with sepsis and septic shock.

#### Correlation with serum lactate

To seek for the correlations between ICU-enterotypes and clinical parameters, we used a subset of 64 samples (one sample per patient) so that the influence of heterogeneity in sample size of patients can be eliminated. These 64 samples were composed of the first collected sample of each patient, whose enterotypes were designated according to enterotype analysis in the former sections. Additionally, the clinical parameters were recorded at the same day with the collection of fecal samples (Table S1). As shown in Table S5, no significant difference was observed in the distribution of sex (*p* = 0.4), age (*p* = 0.68), collecting day of samples (*p* = 0.7323), carbapenem use (*p* = 0.57), breath support (*p* = 0.73) between two ICU-enterotypes, indicating that there was no selection bias of samples between two ICU-enterotypes. We then tested whether crucial clinical parameters, generally used to evaluate the severity of disease condition for septic patients, were different between the two ICU-enterotypes. No significant difference was observed in the SOFA score (*p* = 0.8), APACHE II score (*p* = 0.474), 28-day survival (*p* = 0.36) between the two ICU-enterotypes (Table S5). Interestingly, patients of ICU-enterotype I tended to have higher levels of serum lactate than patients of ICU-enterotype II, though the difference was not significant (2.66 ± 3.30 vs. 1.42 ± 0.52, *p* = 0.07). We then divided the patients into high lactate (serum lactate ≥ 2.5 mmol/L) and low lactate (serum lactate < 2.5 mmol/L) groups. As shown in **Figure 3**A, all of the patients in the high lactate group presented with ICU-enterotype I, and only a certain number of patients in the low lactate group presented with ICU-enterotype II (*p* = 0.007). It turned out that ICU-enterotype I correlated with high serum lactate level.

**Figure 3.**
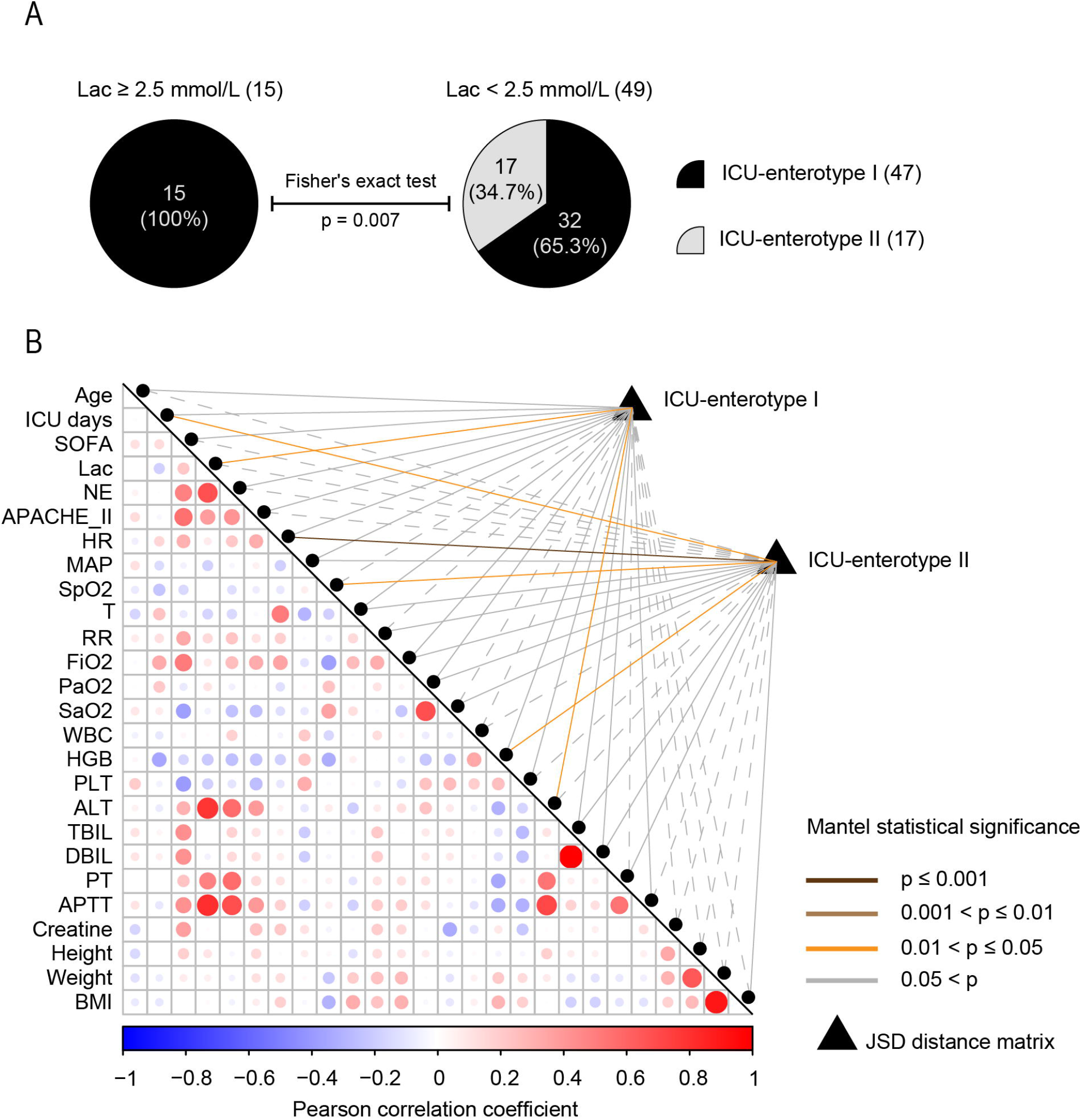
Two ICU-enterotypes are correlated with different clinical parameters. **A**. The pie charts show proportions of patients of two ICU-enterotypes in high lactate group (left panel), as well as low lactate group (right panel). Fisher’s exact test is used to evaluate the statistical significance of the difference in these proportions. **B**. The correlation matrix is produced using Pearson correlation coefficients between each pair of Z-scores-transformed clinical parameters. The size and the color of the circles refer to the strength of these Pearson correlations coefficients, as shown in the bottom of the matrix. The correlations between taxonomic composition of ICU-enterotypes and each of Z-scores-transformed clinical parameters are calculated using Mantel test. Samples of ICU-enterotype I and ICU-enterotype II are tested respectively. The colors of lines refer to the Mantel statistical significance. The solid line refers to positive correlation, while the dashed line refers to negative correlation. The full names of clinical parameters and detailed values of Mantel test are shown in Table S6.

We subsequently performed Mantel test [17] on these 64 samples to test whether clinical parameters of patients can explain gut microbiota variation within ICU-enterotypes. As shown in Figure 3B, the clinical parameters correlated with each other, making them able to cooperatively indicate the severity of illness. Only a few clinical parameters were observed to be able to explain parts of gut microbiota variation (Figure 3B and Table S6. The microbiota variation of ICU-enterotype I was observed to positively correlate with serum lactate concentration (r = 0.148, *p* = 0.0481), suggesting that patients of ICU-enterotype I tended to have more diversified gut microbiota patterns if they have more different serum lactate levels. However, this correlation was not observed in patients with ICU-enterotype II (r = −0.079, *p* = 0.815).

## Discussion

During the period of development of sepsis or septic shock, as well as various medical treatments, ICU patients were observed to contain two quite distinct patterns of gut microbiota, designated as ICU-enterotypes. ICU-enterotypes cannot be explained by the individual host properties such as age, gender and body mass index, as well as external stressors such as infection sites and antibiotic usage. Although the causes of ICU-enterotypes remain unknown, their prevalence, as well as correlation with septic shock and serum lactate level makes them pragmatic for clinical trial, as an independent clinical parameter referring to gut microbiota.

ICU gut microbiota have been previously described as displaying extreme dysbiosis, with loss of health-promoting bacteria and growth of pathogenic bacteria. However, the two ICU-enterotypes identified here indicate that this dysbiosis contains two quite distinct patterns with corresponding sets of loss or growth of bacteria. For example, a loss of *Bacteroides* but a growth of *Enterococcus* was observed in ICU-enterotype II, while the result is diametrically opposite in ICU-enterotype I. Moreover, the bacteria differently distributed between two ICU-enterotypes were observed to partially overlap with the causative pathogens at genus level outside the intestine. Therefore, the dysbiosis of ICU gut microbiota should be analyzed and treated with different strategies according to ICU-enterotypes.

Despite that both ICU-enterotypes are of extreme dysbiosis, their different microbiota patterns correlate with different outcomes. For instance, the larger ratio of *Bacteroides* to *Firmicutes* in ICU-enterotype I, whose ratio > 10 was reported to be correlated with a higher death rate in ICU patients [5], may reflect a critical status of patients. In addition, an unclassified genus of family *Enterobacteriaceae* was observed quite dominant in ICU-enterotype I. Considering the fact that another classified member of this family, named *Enterobacter*, can cause a wide variety of nosocomial infections such as neonatal sepsis with meningitis [18], the identity and potential pathogenicity of this unclassified genus deserve further investigations. On the other hand, *Enterococcus*, which might cause urinary tract infections, abdominal-pelvic infections and endocarditis [19–21], was observed dominant in ICU-enterotype II. Nevertheless, *Enterococcus* was reported to play a role in enhancing immunity and inhibiting overgrowth of opportunistic pathogens, by enhancing the production of short chain fatty acids [22,23] and bacteriocins [24–26]. From this point of view, the overgrowth of *Enterococcus* in ICU-enterotype II might be associated with the lower occurrence of septic shock in ICU-enterotype II through these effects. However, more studies are need to test this speculation. Taken together, both ICU-enterotypes have the potential to lead to sever pathogenic diseases, while the difference in clinical outcome would link to the different patterns of dysbiosis of ICU-enterotypes such as the higher correlation between ICU-enterotype I and septic shock. Moreover, considering the existence of this link, the MHI classifier proposed in this study may facilitate the timely monitoring of ICU-enterotypes variation of patients. The MHI classifier stresses on the improved classification ability of combination of biomarkers, as compared with single biomarker. We believe this strategy can improve the microbiome classification of the other cohort.

ICU-enterotypes can not only serve as an independent clinical parameter for characterization of ICU gut microbiota but also indicate disease severity. In this study, our samples from severe disease conditions showed correlation between ICU-enterotype I and septic shock. In addition, ICU-enterotype I was observed to correlate with high levels of serum lactate, suggesting deterioration of critical hemodynamics. Elevated serum lactate reflects increased anaerobic glycolysis, and is commonly considered to be a marker of tissue hypoxia or hypoperfusion in critically ill patients [27–30]. A certain number of studies suggested that serum lactate > 2mmol/L was independently correlated with mortality in both shock and non-shock patients [30]. Other studies argued that a blood lactate concentration > 0.75mmol/L was independently correlated with increased mortality [31]. In spite of the dispute over the cutoff value, the higher the lactate level, the greater the risk of death, and treatments guided by lactate levels have achieved decreased mortality [32]. Therefore, the ICU-enterotypes have the potential to be the markers of disease severity, thus guiding clinical interventions.

This study also has limitations. Firstly, the lack of healthy subjects and ICU patients without sepsis or septic shock. Secondly, the lack of fecal samples at a certain number of time points restricted us to trace the evolution of ICU-enterotypes in individual patients and to investigate the predictive ability of ICU-enterotypes for developing sepsis or septic shock. Thirdly, the relatively small cohort of our study may have limited the statistical power of the results. Moreover, owing to the lack of detailed clinical information in other studies, we cannot perform a reasonable comparison analysis in other cohorts, though such analysis would undoubtedly be of great interests. Lastly, the comorbidities of the subjects were not considered because of the heterogeneity of the enrolled subjects. Nevertheless, all of the intricate factors such as variety of diseases and medical treatments can be viewed as the causes of the dysbiosis. The impacts of these factors on gut microbiota might be quite different, however, all of these cooperatively resulted in two ICU-enterotypes. This study mainly underscores the existence of two ICU-enterotypes among the cohort of ICU patients with sepsis or septic shock.

## Conclusions

Despite the fact that ICU gut microbiota are of different dysbiosis, two patterns of this dysbiosis are observed quite pervasive among the cohort of ICU patients, designated as ICU-enterotypes. ICU-enterotype I reflects more severe status of septic shock and correlates with deterioration of critical hemodynamics, suggesting higher level of critical treatment. Furthermore, it would be worthwhile to investigate how universal the ICU-enterotypes are among ICU populations around the globe. Better understanding of this can undoubtedly help for more general practical guide for ICU clinical treatment.

## Materials and methods

### Subjects and clinical assessments

This study was conducted in the Critical Care Department of Peking Union Medical College Hospital (PUMCH) of China, which is a tertiary referral hospital with 30 beds in the Critical Care Department. Written informed consent was obtained from all subjects or their legal representatives. Ethical approval was received from the Medical Ethics Committee of PUMCH (ethical reference number HS-1350), and all research was conducted in accordance with the declaration of Helsinki. ICU patients with sepsis or septic shock, who were either directly admitted to the ICU or transferred to the ICU from a hospital ward were enrolled in our study from February 2016 to February 2017. The sequential organ failure assessment (SOFA) score and the acute physiology and chronic health evaluation (APACHE) II score were calculated on a daily basis. Sepsis was defined as when a patient was treated with systemic therapeutic administration of antibiotics due to suspected infection, accompanied by an increase in SOFA score of 2 points or more [33]. Septic shock was identified by a vasopressor requirement to maintain a mean arterial pressure of 65 mmHg or greater and a serum lactate level greater than 2 mmol/L (> 18 mg/dL) in the absence of hypovolemia [33]. Other clinical assessments including physiologic parameters, serum lactate levels and blood tests results were obtained for each patient on a daily basis. Antibiotic usage, the site of infection, the pathogens from positive bacterial cultures, the duration of ICU stay, the occurrence of mechanical ventilation and 28 days survival rate were also collected for each patient.

### Fecal sample collection and 16S rRNA gene sequencing

In this study, the included patients were observed for their first nine consecutive days in ICU (starting from the admission day). Fresh fecal samples were collected from each patient either through defecation or enema for patients with constipation. Enema was performed by the nurses using glycerin enema according to the instructions given by the intensivists. If the patient produced multiple fecal specimens at the same day, only one was collected for test. If the patient was not able to produce fecal specimen on some of the days during the observation duration, then no fecal sample will be available for those days. The fresh fecal samples were then immediately frozen at –80°C. Total bacterial DNA was subsequently extracted from homogenized fecal samples using the QIAamp® Fast DNA Stool Mini kit (QIAGEN, Cat. No. 51604, Germany) according to the manufacturer’s protocol without modification. The V3/V4 hypervariable regions of the 16S ribosomal RNA gene were sequenced using the Illumina MiSeq platform. Mothur [34] was used to merge paired-end reads and trim the reads to meet following quality standard: a quality Phred score ≥ Q20, no ambiguous bases, homo-polymers shorter than 8 bp, and a read length of 300–500 bp. These trimmed reads were then aligned against SILVA 132 reference files (Silva-based alignment of template file for chimera. slayer, release 132) to identify chimeras. Identified chimers were then removed using the UCHIME algorithm [35]. The remaining high-quality sequences were clustered into operational taxonomic units (OTUs) at a 97% sequence identity threshold using UCLUST [36]. The most abundant reads from each of the OTU clusters were taxonomically identified using RDP classifier [37], only accepting annotations with at least 80% confidence. Rarefaction was set at 4000 reads based on curve plateaus for alpha diversity.

### Identification of ICU-enterotypes

The relative abundances of OTUs up to the genus level for each sample were used to perform enterotype analysis [9]. Firstly, the Jensen–Shannon distance (JSD) between each sample was calculated to produce a JSD matrix using relative abundances at genus level. Secondly, the partitioning around medoids (PAM) clustering algorithm was performed on this distance matrix to cluster samples, using the Calinski–Harabasz (CH) index [38] to assess the optimal number of clusters. Thirdly, Silhouette coefficients (SI) [39] were calculated to evaluate the statistical significance of clustering. We set and evaluated the number of clusters from zero to ten, and found two clusters were of the highest CH and SI. The two classified clusters were then named ICU-enterotype I and ICU-enterotype II, respectively. The PAM clustering algorithm, calculation of the CH index and the silhouette index are available in the R packages ‘cluster’ (version 2.0.7) and ‘clusterSim’ (version 0.47-1).

Visualization of the two identified ICU-enterotypes was performed using principal coordinate analysis (PCoA) by the R packages ‘ade4’ (version 1.7-11) and ‘ggplot2’ (version 2.2.1). All fecal samples were projected onto the two-dimensional plane using PCo1 and PCo2 as the most discriminant axes. Shaded ellipses were used to represent the 80% confidence interval, while the dotted ellipse borders indicate the 95% confidence interval, thus wrapping the samples within an enterotype.

### The detection of taxonomic biomarkers of ICU-enterotypes

Taxonomic biomarkers of two identified ICU-enterotypes were firstly picked from the taxonomic abundance table using linear discriminate analysis (LDA) effect size (LEfSe) [40]. These selected biomarkers with LDA scores (log10) > 2.0 were then used as the input for minimum redundancy maximum relevance feature selection (mRMR) [41] to determine the five non-redundant taxonomic biomarkers of each of the two ICU-enterotypes.

### MHI classifier for discriminating ICU-enterotypes

The microbial-based human index (MHI) was employed to determine the combined effects of the taxonomic biomarkers for discriminating ICU-enterotypes. For each sample, ten taxonomic biomarkers, selected by mRMR as described above, were organized according to the formula below to produce the MHI:

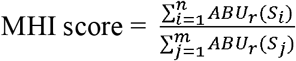

The numerator was the sum of the relative abundance (*ABUr*) of five selected taxonomic biomarkers (Si) of ICU-enterotype I. The denominator was the sum of the relative abundance of five selected biomarkers (Sj) of ICU-enterotype II. The receiver operating characteristic (ROC) curve with 95% confidence intervals and area under the curve (AUC) with 95% confidence intervals were generated for the 131 samples using 9999 stratified bootstrap replicates, to compare the classification ability of the ten individual taxonomic biomarkers and the MHI score.

A threshold was then proposed as the judgment criteria for classifying ICU-enterotypes, i.e., a sample with a MHI score greater than this threshold was classified as ICU-enterotype I and a sample with a MHI score less than this threshold was classified as ICU-enterotype II. The 131 samples were randomly divided into the training set (106 samples, 80%) and the testing set (25 samples, 20%). During the process of training, the threshold was initially set at the min MHI score among samples of training set, and then increased by the value of (the max MHI score – the min MHI score) / 1,000 at every step. This process would terminate when sensitivity and specificity of classification reached to the local optimal value. During the process of testing, the trained threshold was then used to classify the testing set to evaluate the effectiveness of the MHI classifier.

## Data Availability

The 16S rRNA gene sequencing data are available in the Genome Sequence Archive (GSA) section of National Genomics Data Center (project accession number CRA002354). The codes for selection of taxonomic biomarkers and building of MHI classifier were available in GitHub.

https://github.com/HUST-NingKang-Lab/MH-E-I

## Data availability

The 16S rRNA gene sequencing data are available in the Genome Sequence Archive (GSA) section of National Genomics Data Center (project accession number CRA002354). The codes for selection of taxonomic biomarkers and building of MHI classifier were available in GitHub (https://github.com/HUST-NingKang-Lab/MH-E-I).

## Authors’ contributions

YT, KN, YL designed and supervised the study. QH, LS, HH collected samples and recorded clinical information. JL and HF conducted sequencing. WL, MC, PZ, MH performed data analysis. WL, MC wrote the manuscript with input from all authors. YT, KN, YL reviewed and revised the manuscript.

## Competing interests

The authors have declared no competing interests.

## Acknowledgments

This work was partially supported by special Fund for Clinical Research of Wu Jieping Medical Foundation (320.6750.18422), Ministry of Science and Technology of the People’s Republic of China (2018YFC0910502), National Natural Science Foundation of China (31871334 and 31671374). We thank Qiaorong Wang from Nutricia Pharmaceutical Wuxi Co., Ltd for logistic support in the sample collection and sequencing process.

## Supplementary material

### Supplementary figure legends

**Figure S1 The distributions of 131 fecal samples from 64 ICU patients over their first 9 days in ICU since admission**

Each of the rows represents a patient, and each of the columns represents the day. Day 1 also refers to the admission day. The last column represents the mortality observed within 28 days. Only fecal samples at the site of colored circles were obtained and analyzed to calculate enterotypes in this study. The circles represent the characteristics of each patient at each day, whose color represents the ICU-enterotypes and shape (not filled or filled) represents the sepsis or septic shock.

**Figure S2 Distributions of the log-transformed relative abundance of the most dominant genera in two ICU-enterotypes**

The left panel displays the observed distributions using frequency distribution histogram with a density curve. The right panel shows these distributions in ICU-enterotype space. The solid circles refer to samples of septic shock, while the hollow circles refer to samples of sepsis. The most discriminating genera including *Bacteroides* (dominant in ICU-enterotype I, **A**) and *Enterococcus* (dominant in ICU-enterotype II, **B**), as well as their ratio (**C**) show obvious gradient distributions against PCo 1. Another unclassified genus of family *Enterobacteriaceae* (**D**), which is quite dominant in ICU-enterotype I shows obvious gradient distributions against PCo 2.

**Figure S3 Distributions of antibiotic use before/after ICU admission, enema and infection sites of samples in ICU-enterotype space**

The color and shape of circles are based on the ICU-enterotypes and characteristics including the types of antibiotics (**A** and **B**), the way of defecation (**C**), and the infection sites (**D**). In all plots, shaded ellipses represent the 80% confidence interval, while the dotted ellipse borders represent the 95% confidence interval. Boxes represent the interquartile range (IQR) between first and third quartiles and the line inside represents the median. Whiskers denote the lowest and highest values within × IQR from the first and third quartiles, respectively. Statistical significance was tested using the Mann–Whitney–Wilcoxon test, ***P < 0.001, n.s., not significant.

**Figure S4 The comparison of the ability in the classification of ICU-enterotypes between the 10 individual taxonomic biomarkers and the MHI score**

The receiver operating characteristic (ROC) curve with 95% confidence intervals and area under the curve (AUC) with 95% confidence intervals were generated for the 131 fecal samples using 9,999 stratified bootstrap replicates.

### Supplementary table legends

**Table S1 The clinical parameters of 64 ICU patients**

**Table S2 The characteristics of 131 fecal samples**

**Table S3 The statistical evaluation of the number of clusters, based on JSD distance matrix at genus level using 131 fecal samples**

**Table S4 The comparisons of distributions of taxonomic composition between two ICU-enterotypes**

**Table S5 The comparisons of phenotypic characteristics of 64 patients between two ICU-enterotypes**

**Table S6 The correlations between taxonomic composition of first fecal samples of 64 patients and their corresponding clinical parameters**

## Notes

### Clinical Trial

This is a clinical observational study without any medical intervention. The clinical information of the patients were collected retrospectively. This study is approved by the Ethics Committee of the Hospital.

